# SARS-CoV-2 Detection in Sewage in Santiago, Chile - Preliminary results

**DOI:** 10.1101/2020.07.02.20145177

**Authors:** Manuel Ampuero, Santiago Valenzuela, Fernando Valiente-Echeverría, Ricardo Soto-Rifo, Gonzalo P. Barriga, Jonás Chnaiderman, Cecilia Rojas, Sergio Guajardo-Leiva, Beatriz Díez, Aldo Gaggero

## Abstract

The detection of viruses in sewage is a method of environmental surveillance, which allows evaluating the circulation of different viruses in a community. This study presents the first results of sewage surveillance to detect the circulation of SARS-CoV-2 virus in Santiago, Chile. Using ultracentrifugation associated with RT-qPCR, we detected SARS-CoV-2 in untreated and treated wastewater samples obtained two treatment plants, which together process around 85% of the wastewater from the city. This is the first report of detection of SARS-CoV-2 in sewage in Chile and indicates that wastewater surveillance could be a sensitive tool useful as a predictive marker of the circulation of the virus in a population and therefore, be used as an early warning tool.

## The study

On December 31, 2019, the World Health Organization (WHO) reported several cases of pneumonia of unknown etiology detected in the city of Wuhan, located in the province of Hubei, China. The disease was named COVID-19, caused by the SARS-CoV-2 virus. On March 11, 2020 the WHO declared a global pandemic and as of June 22, 2020, there were already 216 countries reporting cases of COVID-19, with a total of 8,860,331 confirmed cases and 465,740 deaths (WHO, 2020) ^1^. The first case of COVID-19 in Chile was detected on March 3, 2020. Since there has been a permanent increase of reported cases and as of June 22 there were 275,648 cases, with a rate of 1,416.6 per 100,000 inhabitants. The largest number of cases are concentrated in the metropolitan region, where the city of Santiago is located, with 220,423 cases and a cumulative incidence rate per 100,000 inhabitants of 2,712.9 ^2^.

Viral detection in wastewater is a method of environmental epidemiological surveillance, which allows evaluating the circulation of different viruses in a community (hepatitis A, hepatitis E, wild and vaccine-derived poliovirus, norovirus, human polyomavirus JC amongst many others). Individuals infected with SARS-CoV-2 have recently been reported to shed the virus in the stool, for prolonged periods of time and with variable viral loads that can reach up to 1 ⨯ 10^6^ copies or more of viral genome/g of fecal material.^3,4,5^ On the other hand, it has been observed that there is a significant percentage of the population carrying the infection asymptomatically and therefore, they are not detected as infected despite these individuals are capable of eliminating the virus through respiratory secretions and feces. Therefore, the circulation of the virus in the population does not become evident until people have moderate or severe symptoms that require hospitalization, or the conditions are in place to carry out a mass diagnosis of the population, allowing early detection of viral infection. In this sense, the detection of SARS-CoV-2 in sewage becomes relevant and there are several worldwide studies that demonstrate the importance of sewage-based surveillance.^6,7,8,9^

We collected 24 hours composite raw sewage samples from two wastewater treatment plants (WWTPs) which together process about 85% of wastewater generated in the city of Santiago (about 8 million inhabitants), i.e. La Farfana and El Trebal. The samples were obtained monthly from March to June 2020, from the influent and effluent of each WWTP in sterile propylene bottles and processed the same day of sampling.

Forthy-two mL of each sample was concentrated by ultracentrifugation and the final pellet was re-suspended in 200 μl of sterile phosphate buffered saline (PBS) pH 7.4 and stored at -80 °C until use.^10,11^

Viral RNA was extracted from concentrated sewage using the QIAamp® Viral RNA Mini kit (QIAGEN, CA, USA) as described by the manufacturer. All samples were analyzed directly and also diluted 1:10 to discard the eventually effects of inhibitors. For detection SARS-CoV-2 RNA we used a TaqMan 2019-nCoV Assay Kit v1 (ThermoFisher Scientific, USA). This kit contains a set of TaqMan RT-PCR assays for detection SARS-CoV-2 RNA and includes three assays that target SARS-CoV-2 genes (ORF1ab, S, and N) and one positive control assay that targets the human RNase P gene.

Our analysis revealed that SARS-CoV-2 RNA was not detected during March and April 2020, in any of the samples tested. This was probably due to the low number of cases in Santiago, which did not allow the virus to be detected in the samples collected from both treatment plants (Figure 1 and Tables 1 and 2). In contrast, the samples obtained in May and June were positive to SARS-CoV-2 RNA, that presented mean cycle threshold (Ct) values ranging from 28,1 to 37,7 (Figure 2). In both WWTP’s, SARS-CoV-2 genome copy numbers progressively increased from May to June, which correlated with the increase in the number of confirmed cases in the city (Figure 3 and Tables 1 and 2). All positive samples had at least two positive reactions, from diluted or undiluted samples.

**Table 1.**
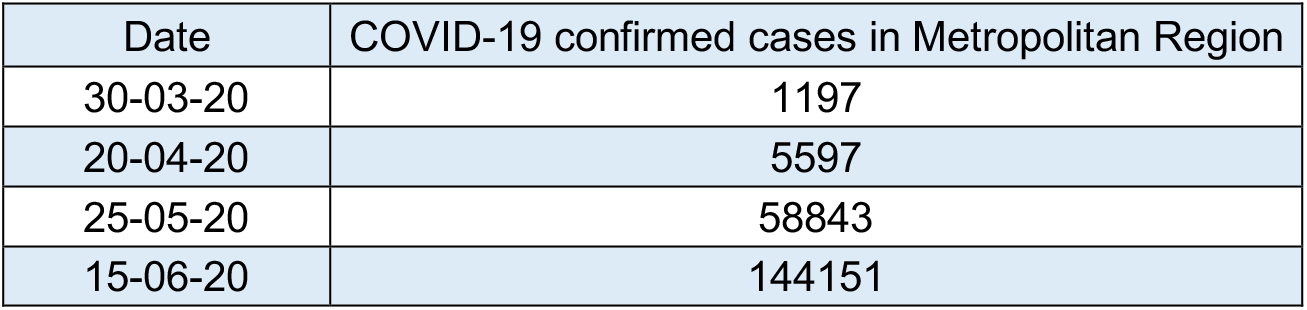
Number of confirmed of cases of COVID-19 in Metropolitan Region between March and June 2020.

| Date | COVID-19 confirmed cases in Metropolitan Region |
| --- | --- |
| 30-03-20 | 1197 |
| 20-04-20 | 5597 |
| 25-05-20 | 58843 |
| 15-06-20 | 144151 |

**Table 2.**
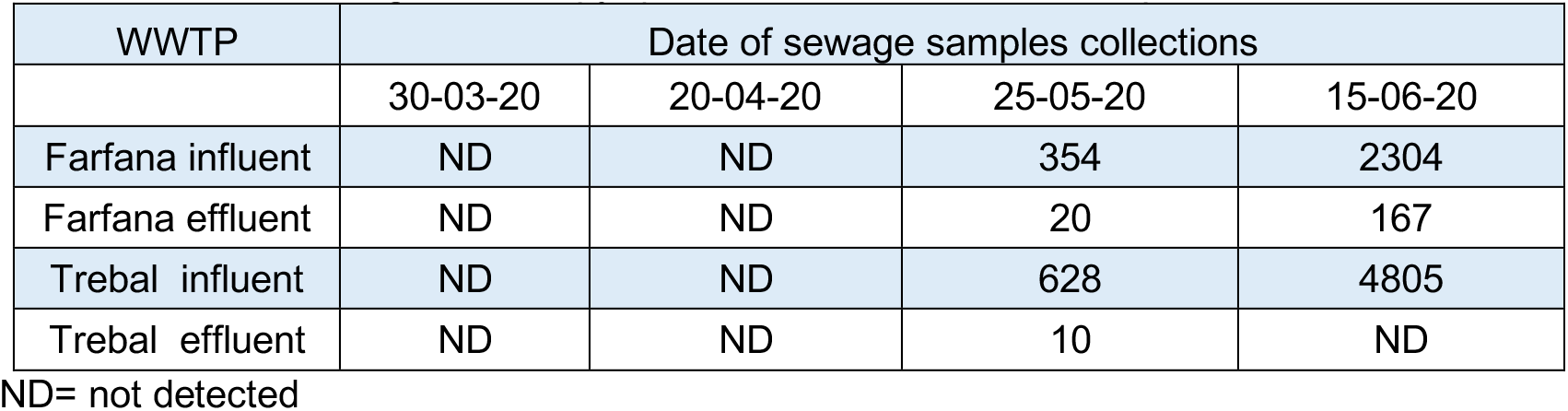
SARS-CoV-2 genome copy quantified in the WWTP’s samples.

**Figure 1.**
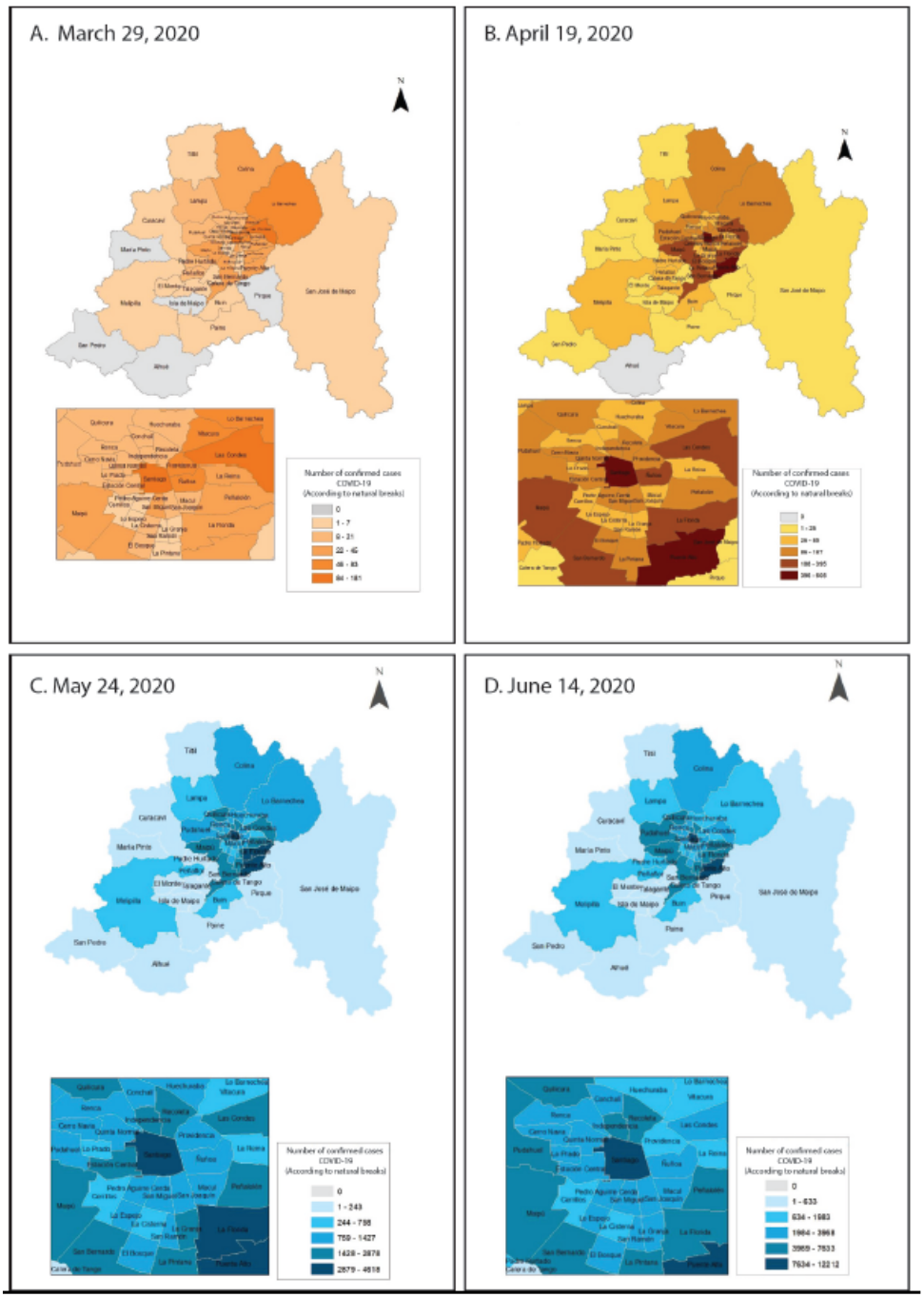
Number of confirmed COVID-19 cases, according to commune of residence in the Metropolitan Region.

**Figure 2.**
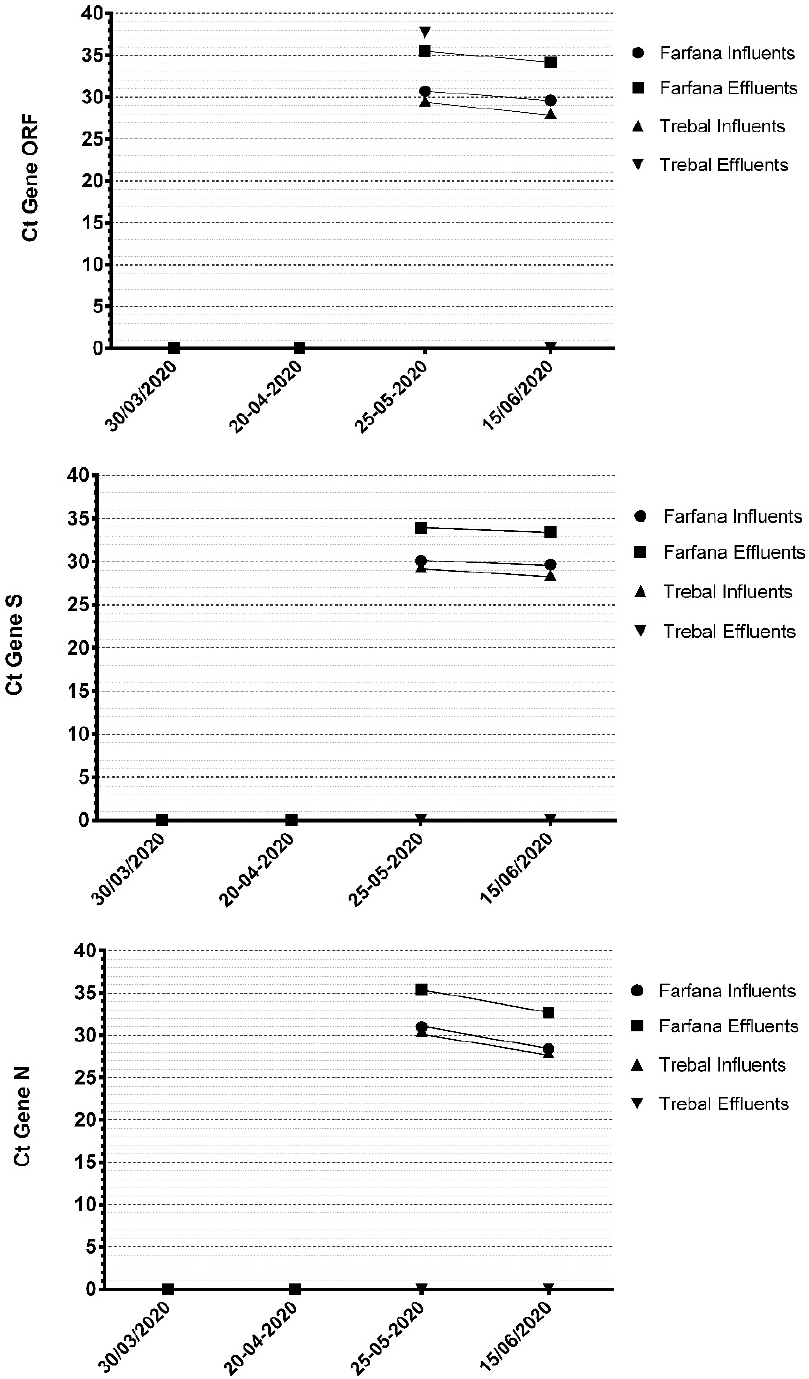
SARS-CoV-2 RNA detection with the mean cycle threshold (Ct) values.

**Figure 3.**
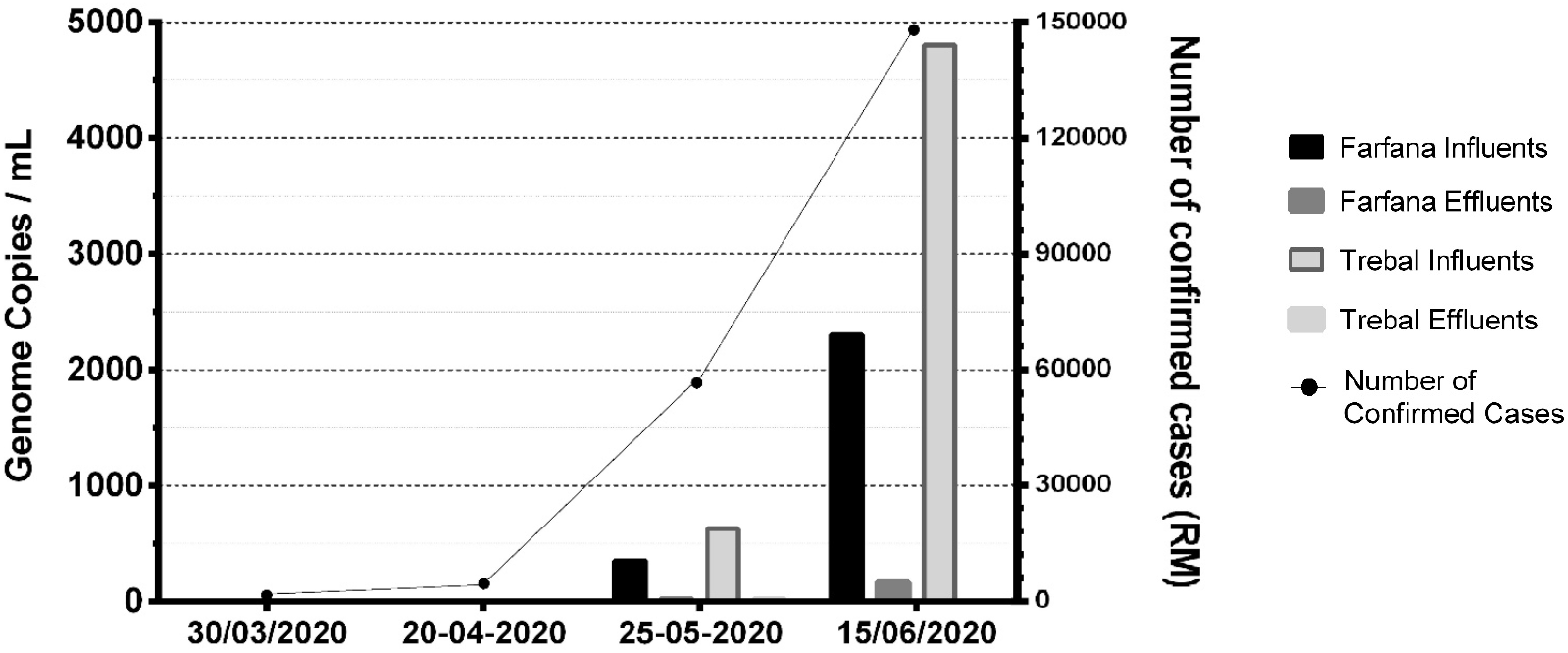
SARS-CoV-2 genome equivalent/mL according to the increased of confirmed cases of COVID-19 per month in Santiago.

In turn, all the samples (March to June) were analyzed for human JC polyomavirus, all of them being positive in ranges of equivalent genome/mL very similar to those previously reported thus, ensuring that the concentration procedure by ultracentrifugation and subsequent nucleic acid extraction was properly performed.^12^

## Conclusions

The goal of this study was to detect SARS-CoV-2 in sewage samples in Santiago, Chile. For this, sewage samples were obtained monthly between March to June 2020 from two WWTPs (La Farfana and El Trebal) which together process about 85% of wastewater generated in Santiago.

In both WWTP’s, the SARS-CoV-2 genome copy numbers progressively increased from May to June, which correlated with the increase in the number of estimated virus shedders in the community.

While enteric transmission of SARS-CoV-2 may be possible and exposure to the virus in sewage could pose a risk to public health, it remains unclear whether the virus is viable under environmental conditions. Our results demonstrated that the search for SARS-CoV-2 in sewage can be used as a predictive marker of the circulation of the virus in the population, including symptomatic and asymptomatic shedders, and therefore can be used as an early warning system as demonstrated years ago for the detection and control of poliovirus worldwide.^13^

## Data Availability

All data referred to in the manuscript are available.

## Acknowledgements

This study was supported by Fondecyt grant n° 1181656.

Authors would like to thank the assistance of the Aguas Andinas WWTP operators, who provided the sewage samples and the Chilean Science, Technology, Knowledge and Innovation Ministry for articulating and coordinating support from the scientific community.

